# A Distinct Nasal Microbiota Signature in Peritoneal Dialysis Patients

**DOI:** 10.1101/2023.02.23.23286379

**Authors:** Iman Khan, Sylvia Wu, Anika Hudson, Clayton Hughes, Gabriel Stryjniak, Lars F. Westblade, Michael J. Satlin, Nicholas Tedrow, Anne-Catrin Uhlemann, Colleen Kraft, Darshana M. Dadhania, Jeffrey Silberzweig, Iwijn De Vlaminck, Carol Li, Vesh Srivatana, John Richard Lee

## Abstract

**Rationale & Objective:** The nasal passages harbor both commensal and pathogenic bacteria. In this study, we sought to characterize the anterior nasal microbiota in PD patients using 16S rRNA gene sequencing.

**Study Design:** Cross-sectional.

**Setting & Participants:** We recruited 32 PD patients, 37 kidney transplant (KTx) recipients, 22 living donor/healthy control (HC) participants and collected anterior nasal swabs at a single point in time.

**Predictors:** We performed 16S rRNA gene sequencing of the V4-V5 hypervariable region to determine the nasal microbiota.

**Outcomes:** Nasal microbiota profiles were determined at the genus level as well as the amplicon sequencing variant level.

**Analytical Approach:** We compared nasal abundance of common genera among the 3 groups using Wilcoxon rank sum testing with Benjamini-Hochberg adjustment. DESeq2 was also utilized to compare the groups at the ASV levels.

**Results:** In the entire cohort, the most abundant genera in the nasal microbiota included: *Staphylococcus, Corynebacterium, Streptococcus*, and *Anaerococcus*. Correlational analyses revealed a significant inverse relationship between the nasal abundance of *Staphylococcus* and that of *Corynebacterium*. PD patients have a higher nasal abundance of *Streptococcus* than KTx recipients and HC participants. PD patients have a more diverse representation of *Staphylococcus* and *Streptococcus* than KTx recipients and HC participants. PD patients who concurrently have or who developed future *Staphylococcus* peritonitis had a numerically higher nasal abundance of *Staphylococcus* than PD patients who did not develop *Staphylococcus* peritonitis.

**Limitations:** 16S RNA gene sequencing provides taxonomic information to the genus level.

**Conclusions:** We find a distinct nasal microbiota signature in PD patients compared to KTx recipients and HC participants. Given the potential relationship between the nasal pathogenic bacteria and infectious complications, further studies are needed to define the nasal microbiota associated with these infectious complications and to conduct studies on the manipulation of the nasal microbiota to prevent such complications.

## INTRODUCTION

The anterior nasal microbiota is at the interface between the external environment and the nasal passages and contains a combination of commensal and pathogenic bacteria. The most common genera defined in healthy individuals in the Human Microbiome Project are *Staphylococcus, Corynebacterium, Propionibacterium*, and *Moraxella* (1). Subsequent studies on the nasal microbiota have revealed microbiota dysbiosis in diseased states such as chronic rhinosinusitis (2) and have linked the nasal microbiota to infectious complications after elective surgical procedures (3).

Peritoneal dialysis (PD) patients undergo dialysis through PD catheter through their abdomen. Despite being taught sterile technique, PD patients experience both exit site infections around the catheter and infectious peritonitis. Prior work has established that pathogenic bacteria in the nasal passages may be associated with infectious complications in PD patients. Luzar et al. reported that *Staphylococcus aureus* nasal colonization was associated with exit site infections in a cohort of 140 PD patients (4). Other studies have found that persistent nasal colonization with *S. aureus* was also associated with peritonitis (5, 6). Decolonization with mupirocin has been suggested to prevent infections and the MUPIROCIN Study Group found that nasal mupirocin prevented *S. aureus* exit site infection (7). Despite these data, International Society of Peritoneal Dialysis (ISPD) guidelines do not support the routine use of nasal mupirocin (8).

Because no study to date has comprehensively evaluated the anterior nasal microbiota in PD patients, we performed a pilot study to evaluate the anterior nasal microbiota using 16S Rrna gene sequencing of the V4-V5 hypervariable region in PD patients, in kidney transplant recipients, and healthy controls.

## METHODS

### Study Cohort Recruitment and Nasal Swab Specimen Collection

From August 2021 to January 2022, we recruited patients receiving peritoneal dialysis (PD), kidney transplant (KTx) recipients, and living donor/healthy control (HC) participants for anterior nasal swab specimen collection. All kidney transplant recipients and living donor candidates were recruited from the clinic. Most PD patients were recruited in the PD clinic; several were recruited during hospitalization. The Weill Cornell Institutional Review Board approved this protocol (IRB # 1604017181) and all participants provided written informed consent.

Anterior nasal swab specimens were collected once from each participant using the Human Microbiome Project protocol. A Copan Eswab (Copan Diagnostics, Murietta, CA, USA) was inserted into the anterior part of one nostril of the participant and turned twice and was then inserted into the anterior part of the other nostril and turned twice. The Copan Eswab was then placed into 1 mL of liquid Amies provided by the Copan Eswab technology and immediately stored on ice or 4°C. Aliquots of 300 uL were created in 2 mL cryovial and stored at -80°C within 12 hours.

### 16S rRNA gene sequencing of the V4-V5 hypervariable region

A single aliquot of approximately 285 μL was deposited into a Qiagen PowerBead glass 0.1 mm tube. Using a Promega Maxwell RSC PureFood GMO and Authentication Kit (AS1600), 1mL of CTAB buffer & 20 μL of RNAse A Solution was added to the PowerBead tube containing the sample. The sample/buffer was mixed for 10 seconds on a Vortex Genie2 and then incubated at 95°C for 5 minutes on an Eppendorf ThermoMixer F2.0, shaking at 1500 rpm. The tube was removed and clipped to a horizontal microtube attachment on a Vortex Genie2 (SI-H524) and vortexed at high-speed for 20 minutes. The sample was removed from the Vortex and centrifuged on an Eppendorf Centrifuge 5430R at 40°C, 12700 rpm for 10 minutes. Upon completion, the sample was centrifuged again for an additional 10 minutes to eliminate foam. The tube was then added to a Promega MaxPrep Liquid Handler tube rack. The Liquid Handler instrument was loaded with proteinase K tubes, lysis buffer, elution buffer, 1000mL tips, 50mL tips, 96-sample deep-well plate, and Promega Maxwell RSC 48 plunger tips. The Promega MaxPrep Liquid Handler instrument was programed to use 300 μL of sample and transfer all sample lysate into Promega Maxwell RSC 48 extraction cartridge for DNA extraction. Upon completion, the extraction cartridge was loaded into Promega Maxwell RSC 48 for DNA extraction & elution. DNA was eluted in 100 μL and transferred to a standard 96-well plate. DNA was quantified using Quant-iT dsDNA High Sensitivity Assay Kit using Promega GloMax plate reader on a microplate (655087). 16S rRNA library generation followed the protocol from the Earth Microbiome Project.

Amplicon libraries were washed using Beckman Coulter AMPure XP magnetic beads. Library quality & size verification was performed using PerkinElmer LabChip GXII instrument with DNA 1K Reagent Kit (CLS760673). Library concentrations were quantified using Quant-iT dsDNA High Sensitivity Assay Kit using Promega GloMax plate reader on a microplate (655087). Library molarity was calculated based on library peak size & concentration. Libraries were normalized to 2nM using the PerkinElmer Zephyr G3 NGS Workstation (133750) and pooled together using the same volume across all normalized libraries into a 1.5mL Eppendorf DNA tube (022431021). Sequencing was performed on an Illumina MiSeq instrument at loading concentration of 7 pM with 15% PhiX, paired-end 250 using MiSeq Reagent Kit v2, 500-cycles (MS-102-2003).

### Bioinformatics Pipeline

Demultiplexed raw reads were processed using the Nextflow (9) nf-core (10) ampliseq pipeline (11), version 2.2.0, with the following parameters: -profile singularity --input SampleSheet.tsv -- FW_primer GTGYCAGCMGCCGCGGTAA --RV_primer CCGYCAATTYMTTTRAGTTT -- metadata Metadata.tsv --outdir results --dada_ref_taxonomy silva --ignore_empty_input_files -- ignore_failed_trimming --min_frequency 10 --retain_untrimmed --trunclenf 240 --trunclenr 160. Specifically, reads were trimmed with cutadapt (12), PhiX and quality filtering, read pair merging, and amplicon sequence variant resolution was performed with DADA2 (13). Subsequent taxonomic assignment was also performed with DADA2, using the Silva reference database (14), version 138. Sequences that were assigned the families, Chloroplast and Mitochondria, were removed from downstream analyses.

### Biostatistical Analyses

The distribution of categorial variables were compared using Fisher’s exact tests. The distribution of continuous variables were compared using Wilcoxon rank sum tests and to account for the comparison of multiple taxa, adjusted p values were calculated using Benjamini-Hochberg adjustment for multiple comparisons. DESeq2 was utilized to detect differences at the ASV between the groups using Benjamini-Hochberg adjustment. Comparison of correlations using a correlational matrix was adjusted for multiple comparisons using the Bonferonni method. All statistical tests were performed using R 4.1.3 in RStudio.

### Data Availability

Sequencing data that support the findings of this study will be made available in the database of Genotypes and Phenotypes (dbGaP) phs002251.v1.p1 after peer-reviewed acceptance. Local institutional review board approval will be needed to access the data.

## RESULTS

### Characteristics of the Study Cohort and Nasal Microbial Sequencing

The microbiota in the anterior nares was performing using 16S rRNA gene sequencing of the V4-V5 hypervariable region in 32 PD patients, 37 KTx recipients, 22 HC participants, and 3 negative controls. A total of 1,116,291 reads with assigned taxonomy was obtained in the cohort of 91 participants with a median of 12,713 assigned reads with an interquartile range of 7,132 and 16,018 assigned reads. The number of assigned reads in the 3 negative controls were 146, 308, and 529, below the number in the cohort of participants.

Table 1 shows the demographics of the participants. In general, the PD patients were older than the HC participants and similar in age to the KTx recipients. More than 50% of PD patients performed automated peritoneal dialysis and 15% had current *Staphylococcus* peritonitis or developed future *Staphylococcus* peritonitis within 10 to 12 months from the nasal specimen collection (last follow up). Approximately a third of KTx recipients received deceased donor transplantation and 32% were on trimethoprim/sulfamethoxazole (TMP-SMX) prophylaxis.

**TABLE 1.** Demographics of the Cohort. Categorical variables are represented by the number followed by the percentage in parentheses. Continuous variables are represented by the median followed by the interquartile range in parentheses.

| Characteristic | PD Cohort (n=32)<br>median abundance | KTx Cohort (n = 37)<br>median abundance | HC Cohort (n = 22)<br>median abundance |
| --- | --- | --- | --- |
| Age, years | 63 (51, 73) | 59 (52, 67) | 51 (33, 61) |
| Female Sex | 20 (63%) | 17 (46%) | 13 (59%) |
| Ethnicity |  |  |  |
| Hispanic | 3 (9%) | 2 (5%) | 3 (14%) |
| Non-Hispanic | 27 (84%) | 33 (89%) | 16 (73%) |
| Declined | 2 (6%) | 2 (5%) | 3 (14%) |
| Race |  |  |  |
| Asian | 4 (11%) | 4 (11%) | 1 (5%) |
| Black | 13 (41%) | 10 (27%) | 5 (23%) |
| White | 11 (34%) | 20 (54%) | 11 (50%) |
| Other | 3 (9%) | 2 (5%) | 1 (5%) |
| Declined | 1 (3%) | 1 (3%) | 4 (18%) |
| History of Hypertension | 26 (81%) | 36 (97%) | 2 (9%) |
| History of Diabetes Mellitus | 9 (28%) | 12 (32%) | 0 (0%) |
| Years on PD | 1.2 (0.6, 2.4) |  |  |
| Automated Peritoneal Dialysis | 21 (66%) |  |  |
| Concurrent or Develops Future<br>*Staphylococcus* Peritonitis | 6 (19%) |  |  |
| Decreased Donor Transplantation |  | 14 (38%) |  |
| Days Post Transplantation |  | 364 (51, 1637) |  |
| History of Prior Transplantation |  | 4 (11%) |  |
| Maintenance Immunosuppression |  |  |  |
| Tacro/mycophenolic mofetil |  | 19 (51%) |  |
| Tacro/mycophenolic mofetil/pred |  | 14 (38%) |  |
| Tacro/mycophenolic acid |  | 1 (3%) |  |
| Tacro/mycophenolic acid/pred |  | 3 (8%) |  |
| Trimethoprim/Sulfamethoxazole PPx |  | 12 (32%) |  |

### Anterior Nasal Microbial Diversity Differs Across the Study Cohort

Microbial diversity among the study participants was measured at the ASV level using the Shannon diversity, an index that evaluates the richness and evenness in a community, as well as Chao1, an index that estimates the total number of ASVs in the specimens. Fig. 1A and B show box and whisker plots of these diversity indices and reveal that PD patients had a significantly higher Shannon diversity index and Chao1 diversity index than KTx recipients (P<0.05, Wilcoxon rank sum test) but similar to the HC participants (P>0.05).

**FIGURE 1.**
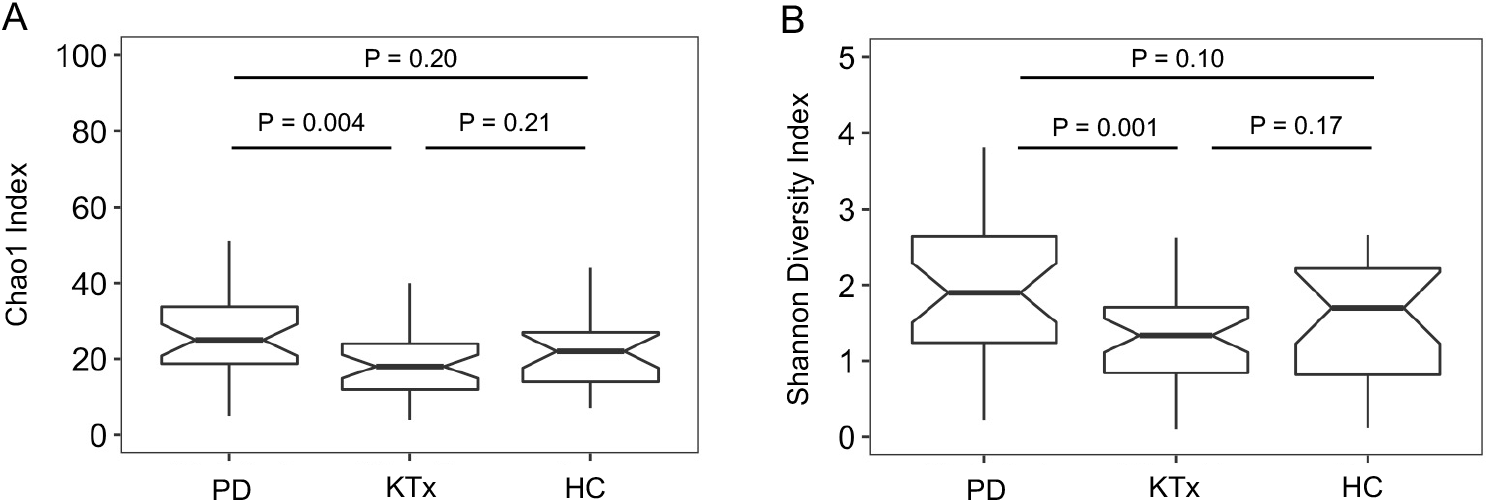
Distinct Differences in Nasal Microbial Diversity among the Study Cohort. **Panel A** shows box and whisker plots of Chao1 index, the estimated number of amplicon sequence variants, in the anterior nasal specimens from the peritoneal dialysis (PD) cohort, the kidney transplant cohort (Ktx), and the living donor /healthy control (HC) cohort. The Chao1 index is on the y axis and the study group is on the x-axis. P value shown was calculated by Wilcoxon rank sum test. **Panel B** shows box and whisker plots of Shannon diversity index, a measure of evenness and richness, in the anterior nasal specimens from the 3 cohorts. The Shannon diversity index is on the y axis and the study group is on the x-axis. P values shown were calculated by Wilcoxon rank sum test.

### *Staphylococcus* Abundance Negatively Correlates with *Corynebacterium* Abundance in Anterior Nasal Specimens

We further evaluated the anterior nasal microbiota among the study cohort at the genus level. At the genus level, the top abundant genera (>1% mean abundance across the cohort) included *Staphylococcus, Corynebacterium, Anaerococcus, Streptococcus*, unspecified *Neisseriaceae, Moraxella, Cutibacterium, Peptoniphilus*, and *Finegoldia* (Fig. 2A). We performed a correlational matrix analysis among each of the genera (Fig. 2B). The relative abundance of *Staphylococcus* was inversely correlated with that of *Corynebacterium* (Pearson r = -0.66, adjusted P value < 0.10, Benjamini-Hochberg adjustment). The relative abundances of *Peptoniphilus* was positively associated with that of *Anaerococcus* (r=0.52, adjusted P value < 0.10) and of *Finegoldia* (r=0.27, adjusted P value < 0.10). The relative abundance of *Finegoldia* was positively associated with that of *Anaerococcus* (r= 0.60, adjusted P value < 0.10). The relative abundance of Unspecified *Neisseriaceae* was positively associated with that of *Cutibacterium* (r= 0.31, adjusted P value < 0.10).

**FIGURE 2.**
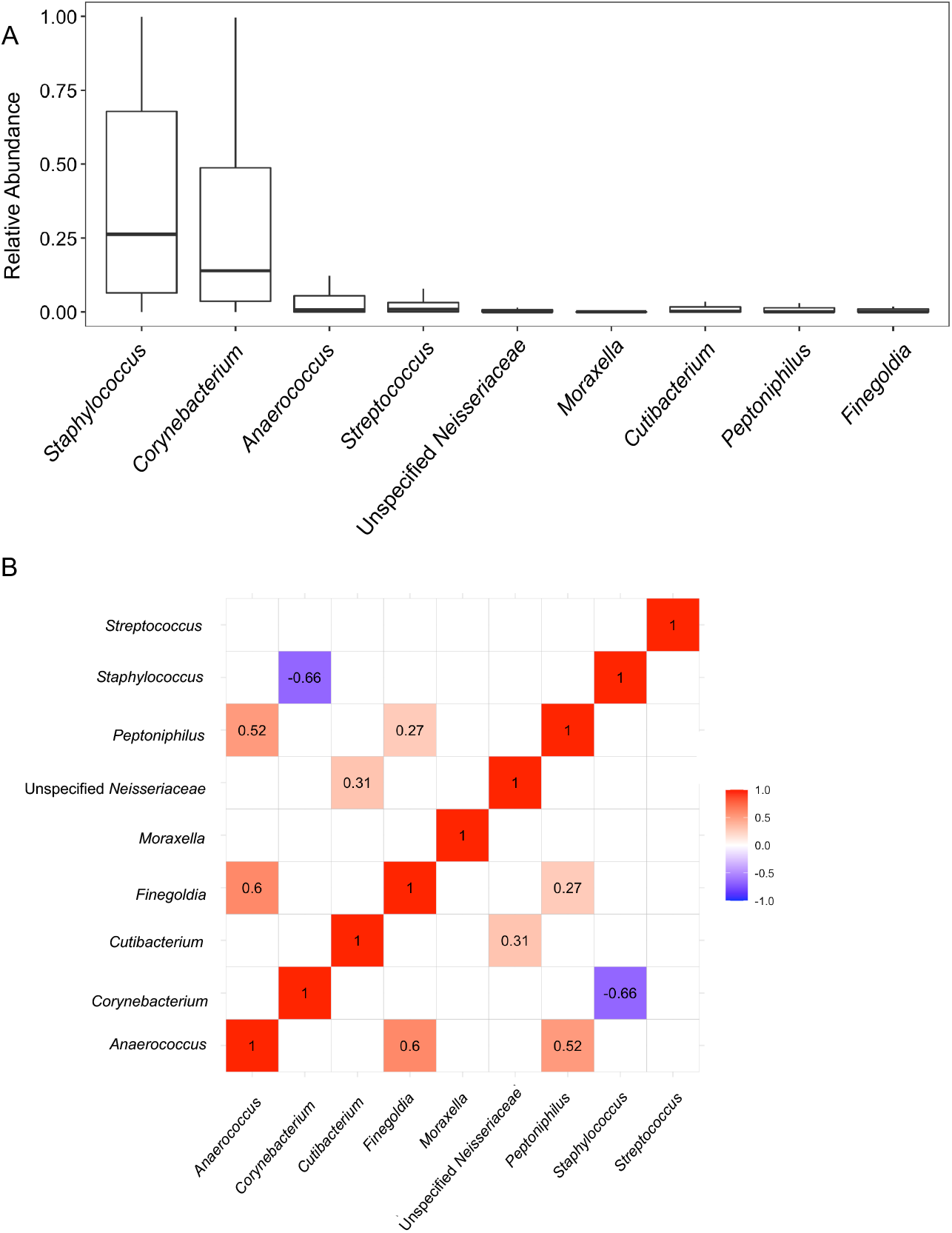
Significant Correlations among the Most Common Genera in the Study Cohort. **Panel A** shows the most common genera in the anterior nasal microbiota (>1% mean relative abundance in the cohort). Box and whisker plots are represented to show the variation in the relative abundance of the genus (y-axis) with the genus on the x-axis. **Panel B** shows a correlational matrix between the nasal abundance of the most common genera using Pearson’s r correlations with Benjamini-Hochberg adjustment for multiple hypotheses. The numbers shown are Pearson’s r correlations that had an adjusted P value < 0.10. The color shows the strength of the correlation with red showing a positive correlation between two genera and blue showing a negative correlation between two genera.

### Distinct Anterior Nasal Microbiota Define PD Patients and Kidney Transplant Recipients

The individual profiles of the top genera in anterior nasal microbiota are shown in the PD patients, KTx recipients, and HC participants (Fig. 3). Fig. 4 shows box and whisker plots of the top 9 taxa among the PD patients, the KTx recipients, and the HC participants and Table 2 shows the comparisons among the groups using Wilcoxon rank sum testing with Benjamini-Hochberg adjustment. PD patients had a distinctly higher relative abundance of *Streptococcus* than KTx recipients or HC participants (Adjusted P value < 0.10, Wilcoxon rank sum test, Benjamini-Hochberg adjustment) (Fig. 4D). PD patients also had lower abundance of unspecified *Neisseriaceae, Moraxella, Cutibacterium*, and *Peptoniphilus* than HC participants (Fig. 4G) (Adjusted P value < 0.10). Kidney transplant recipients had a lower abundance of *Moraxella* than HC participants (Adjusted P value < 0.10). Other than *Streptococcus*, KTx recipients had similar abundance of the top genera compared to PD patients (Adjusted P value > 0.10).

**TABLE 2.**
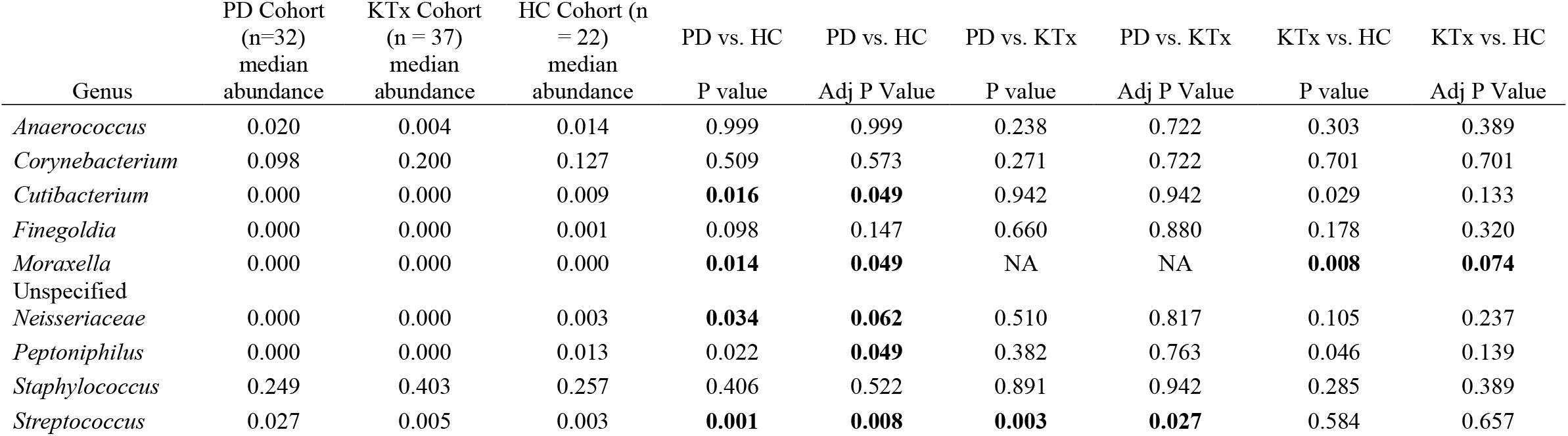
Comparison of the Nasal Abundance Among the 3 Cohorts at the Genus Level. The median abundance of the most common genera are shown for the peritoneal dialysis (PD) cohort, the kidney transplant cohort (KTx), and the living donor /healthy control (HC) cohort. P values shown were calculated using Wilcoxon rank sum test between groups. Adjusted P value (Adj P Value) were calculated using Benjamini-Hochberg adjustment. P values with NA were unable to be calculated because the abundances were 0 in both groups.

**FIGURE 3.**
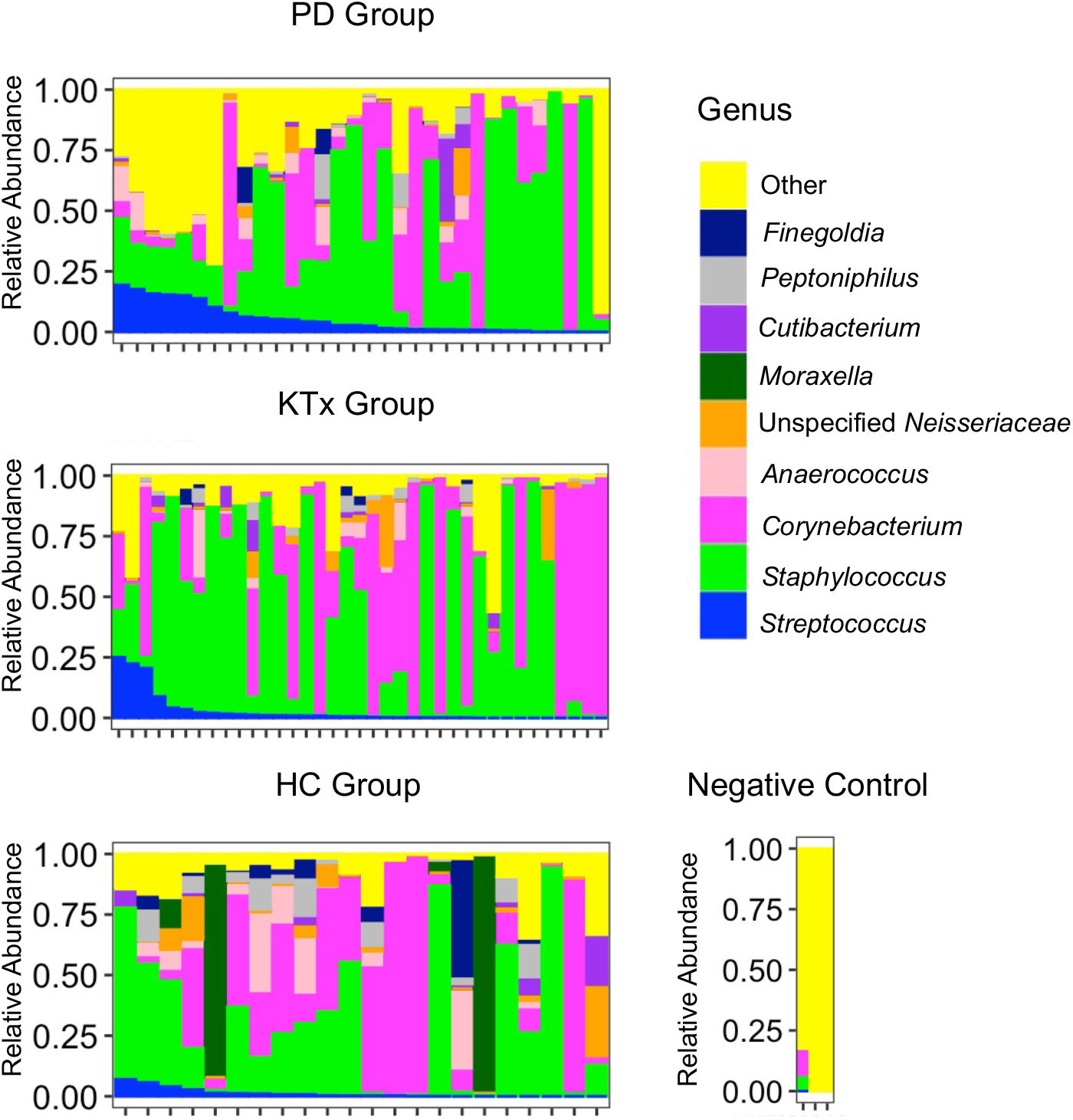
Individual Microbiota Profiles by the Study Cohort. The relative abundance of microbiota is on the y axis and individual nasal specimens are on the x-axis. The relative abundance of each genus is represented by color. The top panel represents anterior nasal microbiota profiles from the 32 peritoneal dialysis (PD) patients, the middle panel represents the anterior nasal microbiota profiles from the 37 kidney transplant (KTx) patients, and the bottom panel represents the anterior nasal microbiota profiles from the 22 potential living donor / healthy control (HC) participants. The right panel represents the microbiota from 3 negative controls.

**FIGURE 4.**
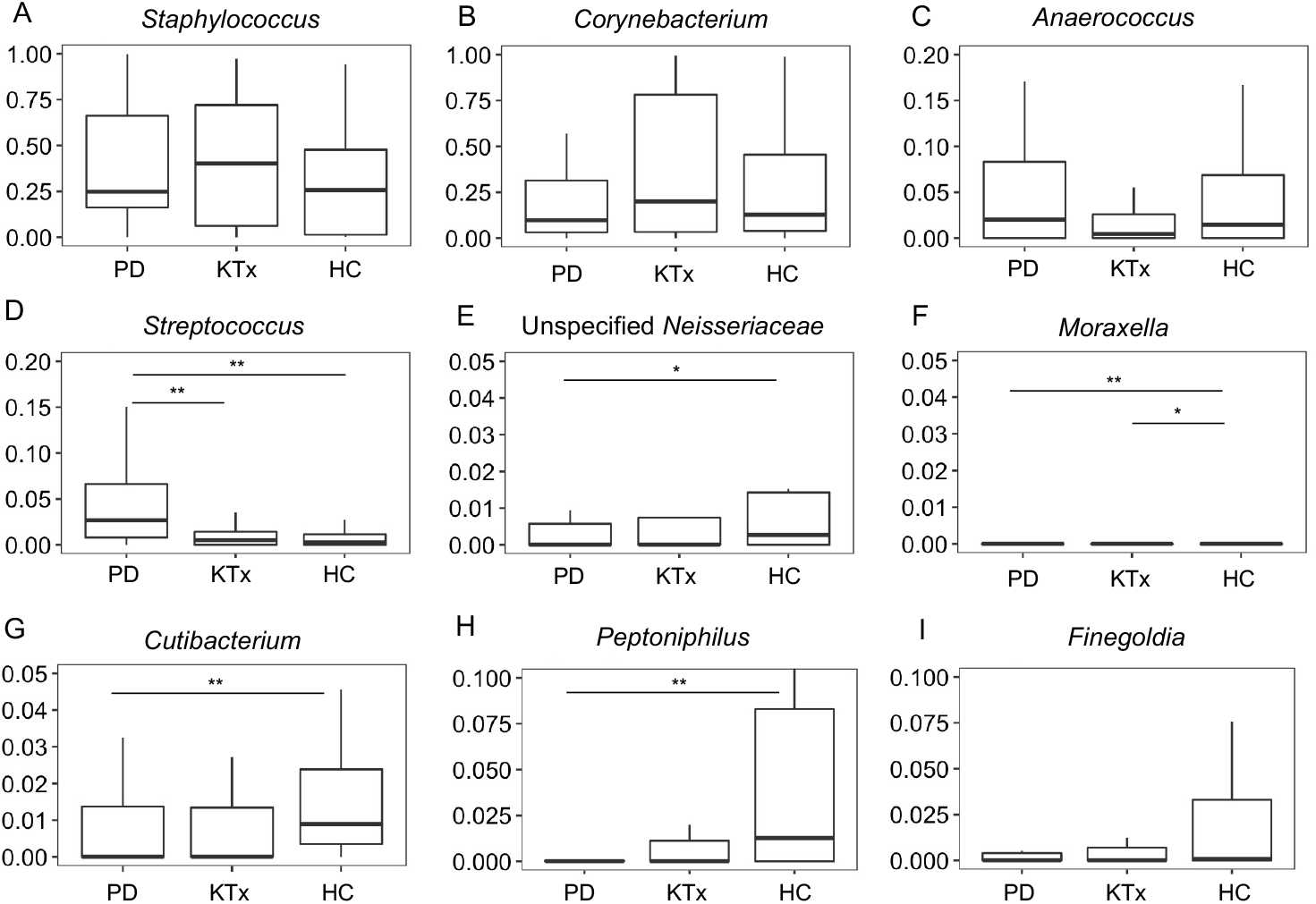
Distinct microbial differences among the cohort at the genus level. Box and whisker plots are represented with the relative abundance of individual genera on the y axis and the group on the x axis. PD, peritoneal dialysis cohort (n=32). KTx, kidney transplant cohort (n=37). HC, living donor / healthy control cohort (n=22). P values were calculated using Wilcoxon rank sum testing with Benjamini-Hochberg adjustment for multiple hypothesis. ** Adjusted P value < 0.05 * Adjusted P value < 0.10. **Panel A**, *Staphylococcus* analysis. **Panel B**, *Corynebacterium* analysis. **Panel C**, *Anaerococcus* analysis. **Panel D**, *Streptococcus* analysis. **Panel E**, Unspecified *Neisseriaceae* analysis. **Panel F**, *Moraxella* analysis. **Panel G**, *Cutibacterium* analysis. **Panel H**, *Peptoniphilus* analysis. **Panel I**. *Finegoldia* analysis.

In order to gain further insight, we evaluated the taxa at the ASV level. We performed pairwise DESeq2 between the groups to identify ASVs that were consistently different among the groups. Fig. 5 shows the significant log2 fold abundance changes between the groups and SI Tables 1 to 3 reveal the changes in the nasal abundances of the groups. Both PD patients and KTx recipients had significantly higher nasal abundances of *Staphylococcus* ASV *#1* and *Corynebacterium* ASV #1 and lower abundance of *Anaerococcus* ASV #1 than HC participants (Adjusted p value < 0.10, Benjamini-Hochberg adjustment). PD patients had higher nasal abundance of *Staphylococcus* ASV #2, *Abiotrophia* ASV #1, and *Porphyromonas* ASV #1 than KTx recipients or HC participants (Adjusted p value < 0.10).

**FIGURE 5.**
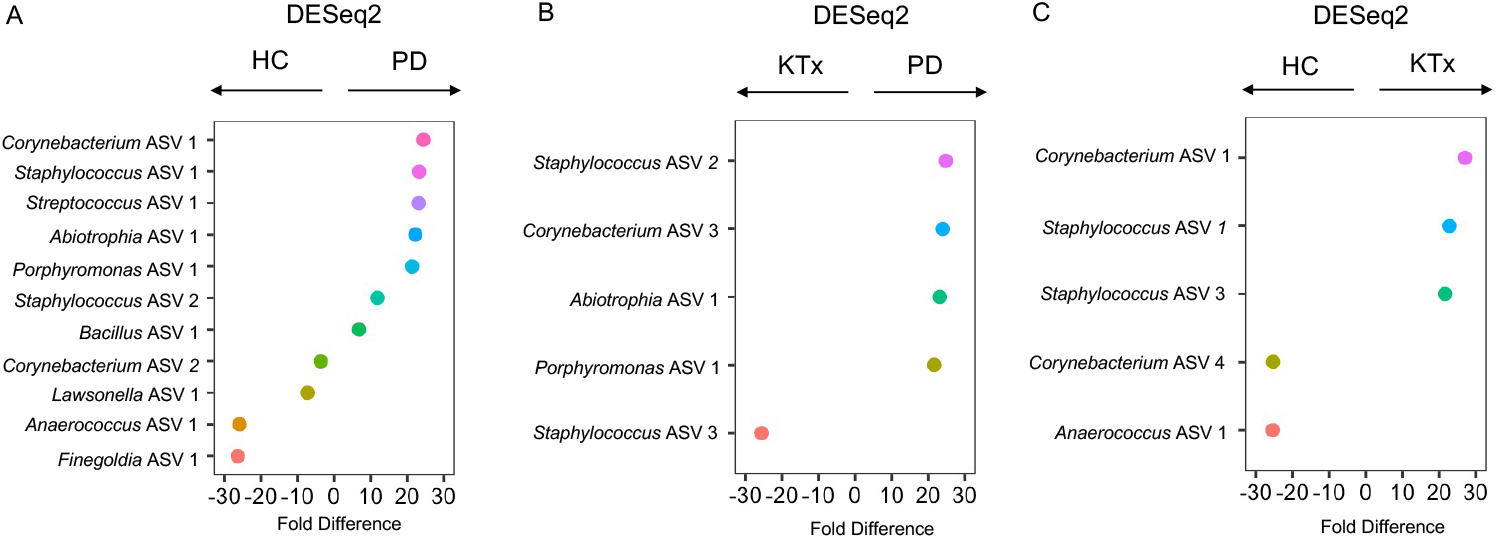
Differential abundance analyses among the cohort at the amplicon sequence variant level. Differential abundance analyses were performed on the anterior nasal microbiota between the groups using DESeq2 with Benjamini-Hochberg adjustment for multiple hypothesis testing. On the y axis is the individual amplicon sequence variant with genus shown and on the x axis is the fold difference in abundance. PD, peritoneal dialysis cohort. KTx, kidney transplant cohort. HC, living donor / healthy control cohort. The fold difference directionality is represented above the graph. **Panel A** represents differential abundance analyses between the HC Group and the PD Group. **Panel B** represents differential abundance analyses between the KTx Group and the PD Group. **Panel C** represents differential abundance analyses between the HC Group and the KTx Group.

### PD Patients Have a More Diverse Representation of Staphylococcus and Streptococcus than KTx Recipients and HC Participants

To further understand why PD patients have higher microbial diversity, we evaluated the diversity of ASVs in the most common genera: *Staphylococcus, Corynebacterium*, and *Streptococcus*. There were 61 different *Staphylococcus* ASVs identified in the whole cohort. PD patients had a significantly higher number of *Staphylococcus* ASVs per specimen than KTx patients (P=0.03, Wilcoxon rank sum test) and HC participants (P=0.04). There were 95 different *Corynebacterium* ASVs identified in the whole cohort. PD patients, KTx patients, and HC participants had similar number of *Corynebacterium* ASVs per specimen (P>0.10). There were 46 different *Streptococcus* ASVs identified in the whole cohort. PD patients had a significantly higher number of *Streptococcus* ASVs per specimen than KTx patients (P=0.04) and HC participants (P=0.05).

### Clinical Factors, Outcomes, and the Nasal Microbiota

We next evaluated the relationship among the nasal microbiota, clinical factors, and outcomes in the cohort. There were no significant differences in the nasal abundances of the most common genera based upon age greater than or equal to 65 years old (SI Table 4). The relative abundance of *Peptoniphilus* was significantly higher in male patients than in female patients (adjusted P value < 0.10) (SI Table 5). Twelve of the kidney transplant recipients were on TMP-SMX prophylaxis for *Pneumocystic jirovecii* prophylaxis and 35 were not. There were no significant differences in the nasal abundance of the most common genera between the kidney transplant recipients on TMP-SMX and those who were not (SI Table 6). In the PD cohort, 6 PD patients concurrently had *Staphylococcus* peritonitis or developed future *Staphylococcus* peritonitis within 10 to 12 months (last follow up) (Staph Peritonitis Group) and 26 PD patients did not (No Staph Peritonitis Group). The nasal abundance of *Staphylococcus* was higher in the Staph Peritonitis Group than in the No Staph Peritonitis Group but the difference was not statistically significant (median abundance 52% vs. 24%, respectively, adjusted P value 0.73). There were no significant differences in the nasal abundance of the other most common genera between the *Staph* Peritonitis Group and the No *Staph* Peritonitis Group (SI Table 7).

## DISCUSSION

This study aimed to describe the anterior nasal microbiota across different groups of patients with kidney disease. We detect a distinct microbial signature in the anterior nares of PD patients compared to KTx recipients and HC participants.

Many of the most common genera in the kidney cohort overlap with those reported in healthy individuals and include *Staphylococccus, Corynebacterium, Finegoldia*, and *Cutibacterium* (15, 16). However, there were some distinct differences among the groups. PD patients had a higher nasal abundance of *Streptococcus* than HC participants or KTx recipients. Interestingly, having a higher nasal abundance of *Streptococcus* has been associated with respiratory infections such as bronchiolitis in infants (17). While the most common type of infectious peritonitis is *Staphylococcus* in origin, *Streptococcus* peritonitis also occurs in PD patients. Our study was not able to directly address whether PD patients with nasal abundance of *Streptococcus* is associated with *Streptococcus* peritonitis and/or respiratory viral infections, but such a link would provide the groundwork for novel approaches to manipulate the nasal microbiota to prevent such complications.

In our analysis, we noticed a higher nasal microbial diversity in the PD patients compared to KTx recipients and HC participants. Further analysis showed that part of this increased microbial diversity may be due to a more diverse representation of *Staphylococcus* and *Streptococcus* in PD patients (Fig. 6). This may have interesting implications as prior data suggests that PD patients with *Staphylococcus aureus* colonization had a higher incidence of exit site infections (Ref). In our study, we did find an increased nasal abundance of *Staphylococcus* in PD patients who had a history of *Staphylococcus* peritonitis and/or developed *Staphylococcus* peritonitis. While the association was not significant, it could be due to the low number of PD patients in the PD Staph peritonitis as our study was not powered to detect the differences. At the 16S rRNA level, we were not able to determine *Staphylococcus* at the species level and this limitation prevented us to assess this association in more detail.

**FIGURE 6.**
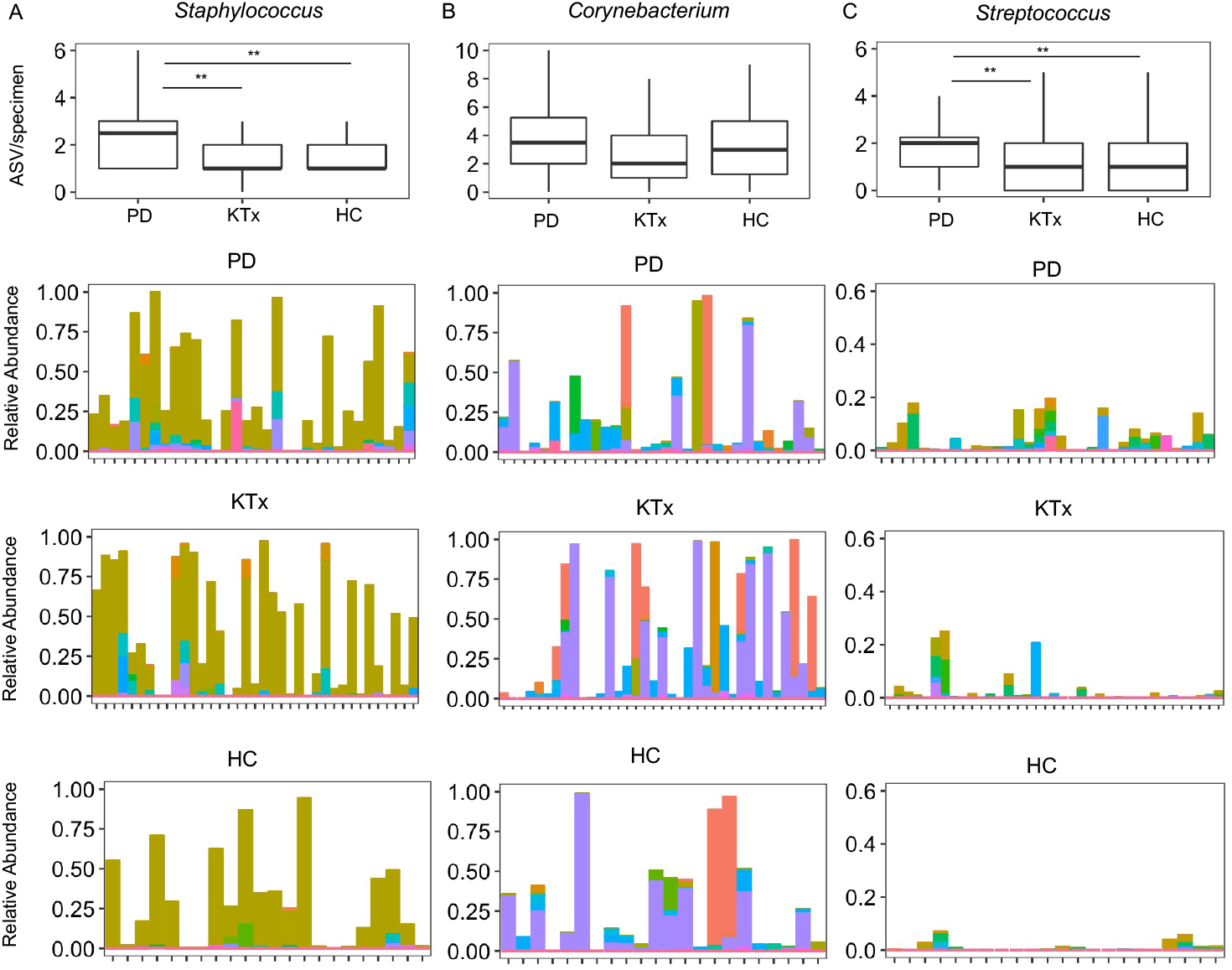
Diverse representation of the most common genera in the study cohort. Each set of graphs represents the number of amplicon sequence variant (ASV) from a particular genus by group. PD, peritoneal dialysis group (n=32). KTx, kidney transplant group (n=37). HC, living donor / healthy control group (n=22). The top graph presents box and whisker plots of the number of ASVs per specimen by group. P value was calculated using Wilcoxon rank sum test: ** P value C< 0.05 * P value < 0.10. The bottom 3 graphs represent box and whisker plots of the relative abundance of individual ASVs from a particular genus. The relative abundance is on the y axis with the color representing individual ASVs and anterior nasal specimens are on the x axis with the second top graph representing the PD Cohort, the second bottom graph representing the KTx Cohort, and the bottom graph representing the HC Cohort. **Panel A** shows the diversity of *Staphylococcus* ASVs in the study cohort. **Panel B** shows the diversity of *Corynebacterium* ASVs in the study cohort. **Panel C** shows the diversity of *Streptococcus* ASVs in the study cohort.

Our data also highlight a strong inverse association between the nasal abundance of *Staphylococcus* and that of *Corynebacterium*. Interesting mechanistic studies have shown a complicated relationship between these two taxa, which represent the most common taxa in the nasal microbiota. One study found that *Corynebacterium* species can secrete antimicrobial peptides against *Staphylococcus aureus (18)*. Another study has shown that *Corynebacterium* species can decrease the virulence of *S. aureus (19)*. Taken together, our data are consistent with the inverse relationship and suggest potential novel approaches to manipulate the nasal microbiota. For example, since *Staphylococcus* peritonitis is much more common than *Corynebacterium* peritonitis, establishing a *Corynebacterium* dominant nasal microbiota may be preventative of *Staphylococcus* in the nasal passages and possibly decrease the risk for *Staphylococcus* exit site infection and/or peritonitis.

A surprising result is that we did not find an association between the TMP/SMX and nasal microbiota differences. TMP/SMX has broad coverage against gram positive cocci including *Staphylococcus* species. There are few studies which have investigated the role of oral antibiotics on the nasal microbiota and it is possible that intra-nasal antibiotics rather than oral antibiotics may more efficiently impact the nasal microbiota. While our study is limited by the population size and the cross-sectional nature, our study raises this possibility.

There are several limitations to our study. As mentioned prior, we are unable to assess species level identification via 16S rRNA gene sequencing of the V4-V5 hypervariable region. Future studies using whole gene 16S rRNA gene sequencing or metagenomic sequencing may provide better resolution on the intricate intra-species competition between the microbiota, particularly between *Staphylococcus* species. and *Corynebacterium* species. Given the low biomass of the nasal microbiota, environmental contamination and/or contamination through the DNA processing steps could artificially introduce microbiota in our specimens. However, we did sequence negative controls (Fig. 3) and the most abundant microbiota identified were not the most common nasal microbiota flora previously reported, suggesting that the nasal microbiota identified in our cohort was present in higher quantities and distinct. The cross-sectional nature of our study provides a snapshot of the microbiota across different groups of patients with kidney disease but does not provide longitudinal changes. Such a longitudinal study may provide more insight into the relationship between the microbiota and clinical factors and outcomes in the populations.

In conclusion, we provide the first description of a distinct nasal microbiota signature in PD patients compared to KTx recipients and HC participants. We find a higher abundance of *Streptococcus* and a more diverse representation of *Staphylococcus* and *Streptococcus* in PD patients. Given the potential relationship between the nasal bacteria and infectious complications in PD patients, further studies are needed to define the nasal microbiota associated with these infectious complications and to conduct studies on the manipulation of the nasal microbiota to prevent such complications.

## Supporting information

STOBE checklist

## Acknowledgments

We thank Dr. Manikkam Suthanthiran for his overall guidance in this project.

---

**SI Table 1.** DESeq2 Analysis of the Nasal Microbiota between the PD Group and the HC Group. The listed ASV were determined to be significantly different using an adjusted p value of 0.10. Base Mean, mean of normalized counts for all samples. Wald stat, Wald statistic. Positive log2 Fold Change is higher in the PD Group.

| Genus | ASV | Base Mean | log2 Fold Change | Standard error | Wald stat | P value | Adj P value |
| --- | --- | --- | --- | --- | --- | --- | --- |
| *Finegoldia* ASV #1 | 1f016ec26e4774c86f029301b556de28 | 24.3 | -26.4 | 2.9 | -9.0 | 3.4E-19 | 1.4E-17 |
| *Anaerococcus* ASV #1 | ae7cf0f32080cf5e62b3fbd903a997af | 16.5 | -25.9 | 3.0 | -8.7 | 2.2E-18 | 4.4E-17 |
| *Lawsonella* ASV #1 | b89dbfe5c74375ff3d4922b41c470740 | 49.0 | -7.3 | 2.9 | -2.5 | 1.3E-02 | 5.1E-02 |
| *Corynebacterium* ASV #2 | c91b64f1467982a81922d331a310f5dc | 1172.2 | -3.7 | 1.6 | -2.2 | 2.5E-02 | 9.2E-02 |
| *Bacillus* ASV #1 | 5565e52f91bc244013249656795409ca | 18.7 | 6.9 | 1.8 | 3.8 | 1.7E-04 | 7.7E-04 |
| *Staphylococcus* ASV #2 | b273d7d05f0525bb87418b0138d90605 | 86.0 | 11.8 | 2.2 | 5.3 | 1.2E-07 | 6.1E-07 |
| *Porphyromonas* ASV #1 | 969732ae717bf5a9282f4781499aadb4 | 10.0 | 21.4 | 3.0 | 7.2 | 6.3E-13 | 3.6E-12 |
| *Abiotrophia* ASV #1 | afce1dc2c06acb90ad5d3da0fffbcd02 | 9.7 | 22.3 | 2.8 | 7.9 | 3.5E-15 | 2.4E-14 |
| *Streptococcus* ASV #1 | 4481bc66d3a54f79e6abb8b557a2a104 | 19.3 | 23.2 | 2.9 | 7.9 | 2.5E-15 | 2.4E-14 |
| *Staphylococcus* ASV #1 | 7b06c22a4ff85c9b2cbbf57a53462e13 | 41.9 | 23.4 | 3.0 | 7.9 | 3.6E-15 | 2.4E-14 |
| *Corynebacterium* ASV #1 | 0560198302010f9b110d4ec9897e14d0 | 45.9 | 24.5 | 3.0 | 8.2 | 1.8E-16 | 2.3E-15 |

**SI Table 2.** DESeq2 Analysis of the Nasal Microbiota between the PD Group and the KTx Group. The listed ASV were determined to be significantly different using an adjusted p value of 0.10. Base Mean, mean of normalized counts for all samples. Wald stat, Wald statistic. Positive log2 Fold Change is higher in the PD Group.

| *Genus* | ASV | Base Mean | log2 Fold Change | Standard error | Wald stat | P value | Adj P value |
| --- | --- | --- | --- | --- | --- | --- | --- |
| *Staphylococcus ASV #3* | 0f89ba63b47f3b624c13293cb4d56486 | 16.5 | -25.6 | 2.9 | -8.8 | 1.9E-18 | 2.0E-16 |
| *Porphyromonas ASV #1* | 969732ae717bf5a9282f4781499aadb4 | 7.4 | 21.6 | 2.9 | 7.4 | 1.1E-13 | 4.8E-12 |
| *Abiotrophia ASV #1* | afce1dc2c06acb90ad5d3da0fffbcd02 | 7.3 | 23.3 | 2.8 | 8.4 | 4.3E-17 | 3.0E-15 |
| *Corynebacterium ASV #3* | 36f00e24a741c19a5ee20f58919004ad | 12.4 | 24.0 | 2.9 | 8.2 | 1.7E-16 | 8.8E-15 |
| *Staphylococcus ASV #2* | b273d7d05f0525bb87418b0138d90605 | 65.8 | 24.8 | 2.4 | 10.2 | 1.5E-24 | 3.1E-22 |

**SI Table 3.** DESeq2 Analysis of the Nasal Microbiota between the KTx Group and the HC Group. The listed ASV were determined to be significantly different using an adjusted p value of 0.10. Base Mean, mean of normalized counts for all samples. Wald stat, Wald statistic. Positive log2 Fold Change is higher in the KTx Group.

| *Genus* | ASV | Base Mean | log2 Fold Change | Standard error | Wald stat | P value | Adj P value |
| --- | --- | --- | --- | --- | --- | --- | --- |
| *Anaerococcus ASV #1* | ae7cf0f32080cf5e62b3fbd903a997af | 12.0 | -25.4 | 3.0 | -8.5 | 2.8E-17 | 2.2E-15 |
| *Corynebacterium ASV #4* | 9f6c00ff9e1f455f42f211770d60dec0 | 11.2 | -25.3 | 3.0 | -8.4 | 3.6E-17 | 2.2E-15 |
| *Staphylococcus ASV #3* | 0f89ba63b47f3b624c13293cb4d56486 | 12.5 | 21.6 | 3.0 | 7.2 | 7.4E-13 | 2.8E-11 |
| *Staphylococcus ASV #1* | 7b06c22a4ff85c9b2cbbf57a53462e13 | 11.8 | 22.9 | 3.0 | 7.6 | 2.8E-14 | 1.3E-12 |
| *Corynebacterium ASV #1* | 0560198302010f9b110d4ec9897e14d0 | 237.4 | 27.1 | 3.0 | 9.0 | 2.2E-19 | 4.1E-17 |

**SI Table 4.** Comparison of Nasal Microbiota Based on Patient’s Age at the Genus Level. P value was calculated between groups using Wilcoxon rank sum test. Adjusted p value (Adj P value) was calculated using Benjamini-Hochberg adjustment.

| Genus | Age $\geq 65$ | Age $< 65$ | P value | Adj P value |
| --- | --- | --- | --- | --- |
|  | n = 28 | n = 63 |  |  |
| *Staphylococcus* | 0.187 | 0.346 | 0.11 | 0.40 |
| *Finegoldia* | 0.000 | 0.001 | 0.11 | 0.40 |
| *Anaerococcus* | 0.001 | 0.013 | 0.14 | 0.40 |
| *Moraxella* | 0.000 | 0.000 | 0.18 | 0.40 |
| *Unspecified Neisseriaceae* | 0.000 | 0.000 | 0.30 | 0.48 |
| *Streptococcus* | 0.012 | 0.007 | 0.32 | 0.48 |
| *Corynebacterium* | 0.124 | 0.139 | 0.56 | 0.72 |
| *Peptoniphilus* | 0.000 | 0.000 | 0.66 | 0.74 |
| *Cutibacterium* | 0.003 | 0.002 | 0.81 | 0.81 |

**SI Table 5.**
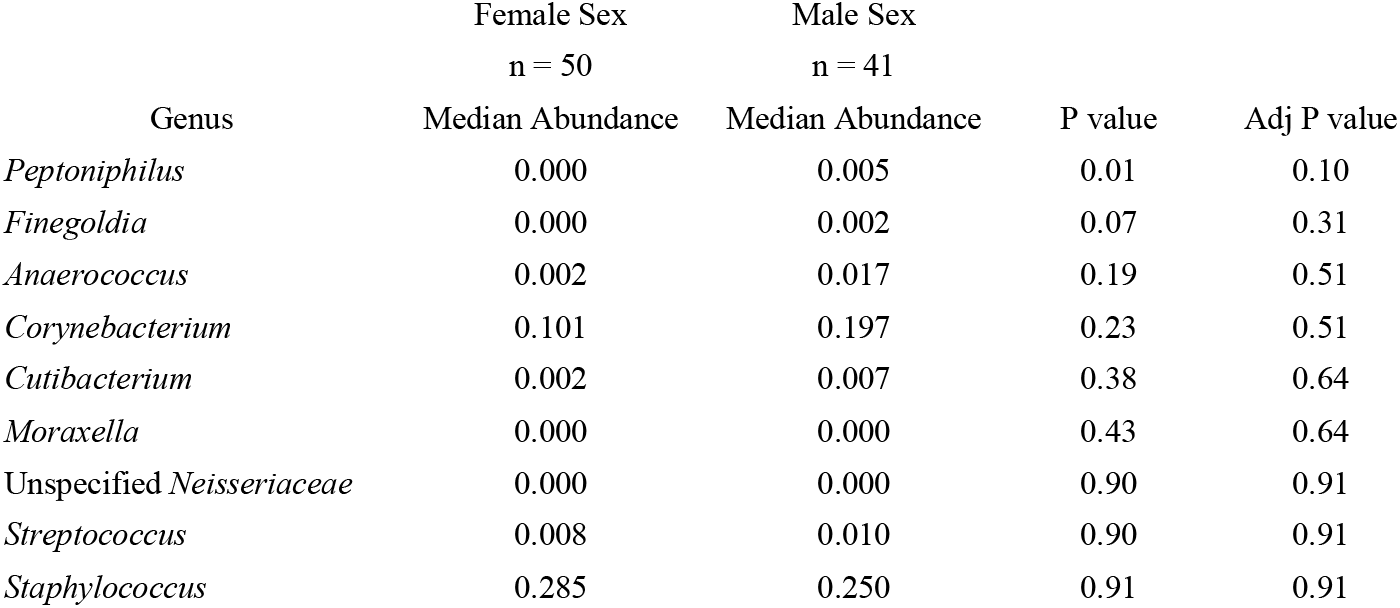
Comparison of Nasal Microbiota Based on Patient’s Sex at the Genus Level. P value was calculated between groups using Wilcoxon rank sum test. Adjusted p value (Adj P value) was calculated using Benjamini-Hochberg adjustment.

**SI Table 6.** Comparison of Nasal Microbiota Based on Trimethoprim/Sulfamethoxazole usage in the Kidney Transplant Recipients at the Genus Level. P value was calculated between groups using Wilcoxon rank sum test. Adjusted p value (Adj P value) was calculated using Benjamini-Hochberg adjustment. P values with NA were unable to be calculated because the abundances were 0 in both groups. KTx, Kidney Transplant; TMP/SMX, trimethoprim/sulfamethoxazole.

| Genus | KTx on TMP/SMX | KTx off TMP/SMX | P value | Adj P value |
| --- | --- | --- | --- | --- |
|  | n = 12 | n = 25 |  |  |
|  | Median Abundance | Median Abundance |  |  |
| *Finegoldia* | 0.000 | 0.002 | 0.02 | 0.15 |
| *Peptoniphilus* | 0.000 | 0.000 | 0.04 | 0.15 |
| *Anaerococcus* | 0.0002 | 0.007 | 0.06 | 0.15 |
| *Corynebacterium* | 0.101 | 0.314 | 0.36 | 0.59 |
| *Cutibacterium* | 0.000 | 0.000 | 0.37 | 0.59 |
| *Staphylococcus* | 0.363 | 0.487 | 0.53 | 0.71 |
| Unspecified *Neisseriaceae* | 0.000 | 0.000 | 0.70 | 0.80 |
| *Streptococcus* | 0.002 | 0.005 | 0.84 | 0.84 |
| *Moraxella* | 0.000 | 0.000 | NA | NA |

**SI Table 7.**
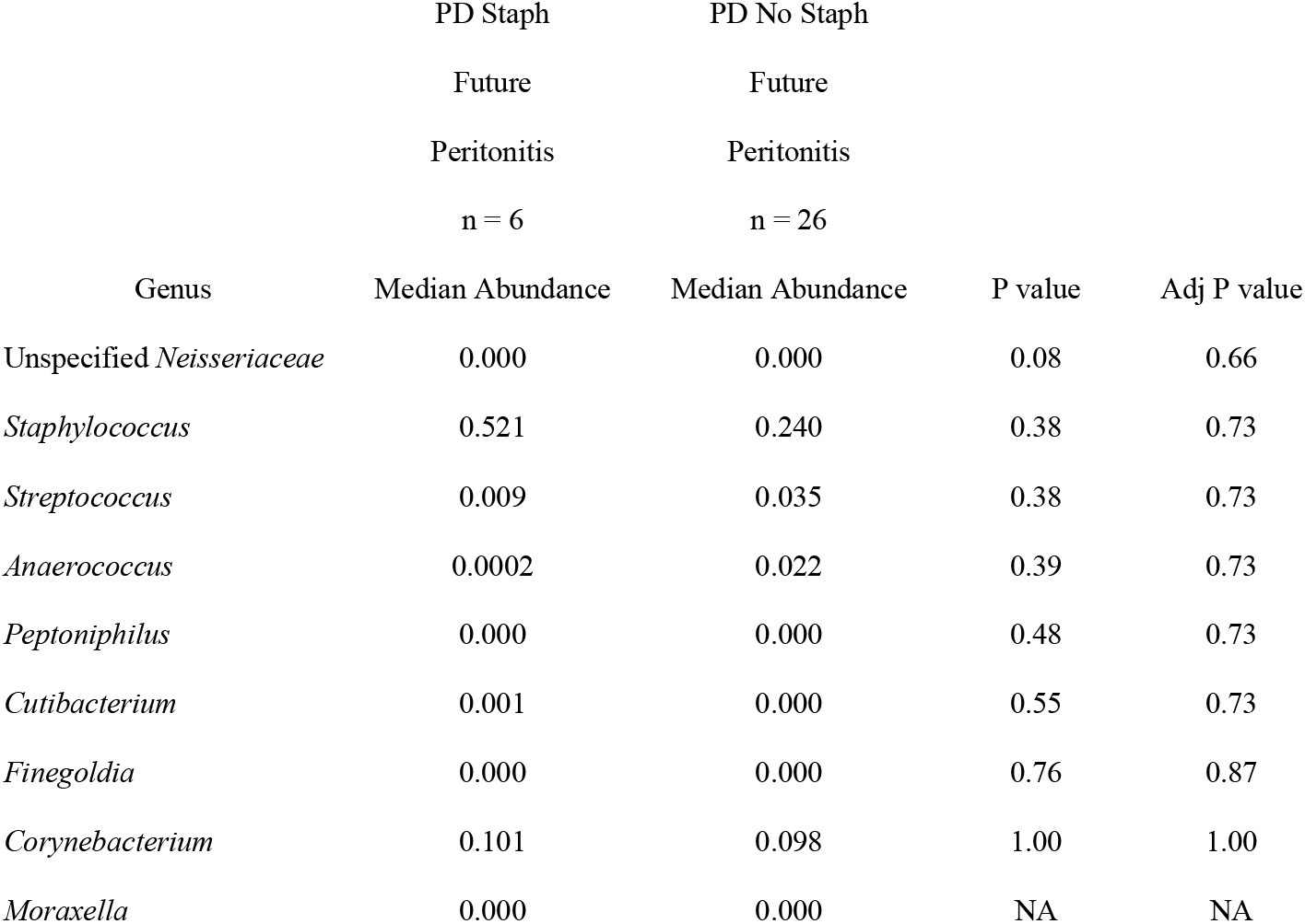
Comparison of Nasal Microbiota Based on PD patients who developed *Staphylococcus* peritonitis status and PD patients who did not develop *Staphylococcus* peritonitis at the Genus Level. P value was calculated between groups using Wilcoxon rank sum test. Adjusted p value (Adj P value) was calculated using Benjamini-Hochberg adjustment. P values with NA were unable to be calculated because the abundances were 0 in both groups. PD Staph peritonitis cohort was defined as PD patients who currently had *Staphylococcus* peritonitis or developed *Staphylococcus* peritonitis withn 10 to 12 months (last follow up).

